# Whole-population perspective is needed for analyses and actions to address linear growth faltering in low- and middle-income countries

**DOI:** 10.1101/2024.06.24.24309409

**Authors:** Daniel E. Roth, Kelly M. Watson, Diego G. Bassani

**Affiliations:** Centre for Global Child Health, The Hospital for Sick Children, Toronto, Canada; Department of Paediatrics, University of Toronto, Toronto, Canada; Dalla Lana School of Public Health, University of Toronto, Toronto, Canada; Department of Nutritional Sciences, University of Toronto, Toronto, Canada

## Abstract

Linear growth faltering (LGF), or slower than normal growth in height, is widely considered an indicator of suboptimal conditions affecting children’s development and health in low- and middle-income countries (LMICs). Recently, Benjamin-Chung and collaborating members of the Healthy Birth, Growth and Development Knowledge integration (HBGDki) consortium described the early onset and low reversal rates of LGF in 32 cohort studies that followed over 52,000 children from birth to 24 months of age in 14 countries. Their adoption and extension of conventionally used growth metrics to describe faltering patterns led to findings that echo a long-standing assumption that LGF in resource-constrained settings occurs mainly during early infancy and is mostly irreversible thereafter. Here, we discuss limitations of their methods and suggest an alternative approach that leads to different conclusions about the rate and timing of LGF in LMICs.

---

Linear growth faltering (LGF), or slower than normal growth in height, is widely considered an indicator of suboptimal conditions affecting children’s development and health in low- and middle-income countries (LMICs). Recently, Benjamin-Chung and collaborating members of the Healthy Birth, Growth and Development Knowledge integration (HBGDki) consortium described the early onset and low reversal rates of LGF in 32 cohort studies that followed over 52,000 children from birth to 24 months of age in 14 countries^1^. Their adoption and extension of conventionally used growth metrics to describe faltering patterns led to findings that echo a long-standing assumption that LGF in resource-constrained settings occurs mainly during early infancy and is mostly irreversible thereafter. Here, we discuss limitations of their methods and suggest an alternative approach that leads to different conclusions about the rate and timing of LGF in LMICs.

Infant LGF in LMICs is a whole-population phenomenon, meaning that even the tallest infants are shorter than they would be under optimal growth conditions. When a population of infants experiences LGF, it leads to a decline in the mean length-for-age z-score (LAZ) and a downward shift of the entire LAZ distribution^2^. A conventional indicator of childhood undernutrition is the proportion of children who are stunted (% with LAZ<-2), which is an acceptable population-level measure of prior LGF because, like mean LAZ, it enables direct comparison of a population’s height distribution to an international norm. However, classifying individual children within that population as stunted is uninformative because stunted children have not generally experienced slower postnatal growth than their non-stunted peers^3^. HBGDki investigators observed whole-population LGF in their LMIC cohorts but described growth patterns based on the proportions of infants who changed stunting status between sequential age windows of observation, primarily using metrics they referred to as incident stunting onset (newly crossing the threshold of LAZ = - 2 from a preceding higher LAZ value) and stunting reversal (crossing the threshold of LAZ = -2 from a preceding lower LAZ value).

We first hypothesized that patterns of stunting onset and reversal reported by HBGDki are intrinsic features of whole-population LGF, similar to stunting prevalence. To test this, we simulated a birth cohort that experienced a downward shifting LAZ distribution similar to that observed by HBGDki in their South Asian (SA) cohorts (**Supplementary Information & Extended Data Table 1**). A key advantage of this simulation is the absence of factors that differentially affect individuals’ growth across the height distribution or at different ages. Conceptually, this means that those with LAZ<-2 at a given age were similar to non-stunted infants with respect to both the extent to which they experienced prior LGF and their subsequent responsiveness to factors that could promote rapid growth. Therefore, in the simulated cohort, child-level changes in LAZ between sequential observations result from only two determinants: whole-population shifting of the LAZ distribution and between-timepoint length/height correlation. Because such correlations are always imperfect (i.e., corr<1), children’s ranks in the LAZ distribution readily change from one timepoint to the next, and the further a child’s length/height is from the group mean at one timepoint, the more likely that child’s measurement will be closer to the group mean at a subsequent timepoint, a statistical phenomenon known as regression towards the mean (RTM)^4^.

Estimates of stunting incidence and reversal in the simulated cohort followed a remarkably similar pattern over time (i.e. age) as those reported by HBGDki (**Figure 1**), providing a robust proof-of-concept that a few basic assumptions about whole-population LGF are sufficient to approximate their real-world observations. This suggests that stunting incidence and reversal patterns reported by HBGDki reflect statistical characteristics of a downward shifting LAZ distribution and, therefore, do not yield new insights into differential age effects of LGF or the unique experiences of children with an observed LAZ below -2 (or any other threshold value, as explained in more detail below). We did not expect our simulated estimates to be identical to those reported by HBGDki because we could not replicate the exact shape and dispersion of the LAZ distributions or between-timepoint correlations and because their published estimates were based on aggregated cohorts with substantial missingness, which could have increased some estimates of incident stunting onset (**Extended Data Table 2**).

**FIGURE 1.**
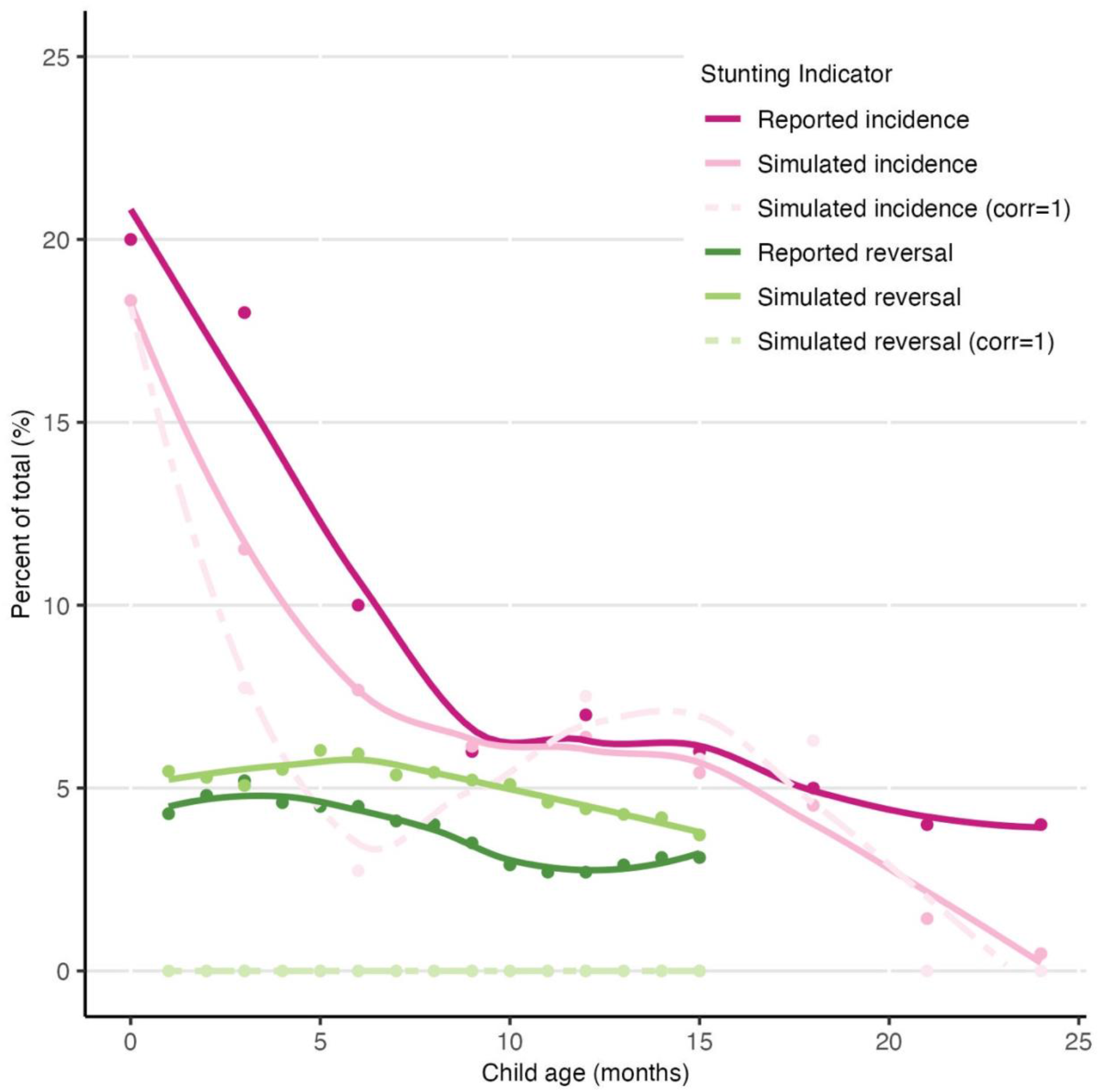
Incident stunting onset and reversal (%) at tri-monthly intervals (onset) and monthly intervals (reversal), reported by Benjamin-Chung et al. (“Reported incidence/reversal”), in a simulated cohort with a realistic between-timepoint correlation matrix (“Simulated incidence/reversal”), and in a simulated cohort with perfect correlation for all intervals (“Simulated incidence/reversal (corr=1)”). Circles denote point estimates and lines represent loess curves for each set of points (bandwidth = 0.75). Simulations and calculation of stunting incidence and reversal are explained in Supplementary Information.

A key implication of whole-population LGF is that the value of the LAZ threshold used to define any prevalence/incidence indicator is irrelevant, which explains why HBGDki observed similar patterns of onset and reversal using the *severe stunting* threshold (LAZ<-3). In the context of whole-population LGF, subgrouping children based on whether they crossed a given LAZ threshold can lead to unsubstantiated conclusions about the causes and consequences of LGF because characteristics associated with *stunting* (i.e., relatively short stature versus other children in the same population) may be misinterpreted as risk factors for LGF (i.e., slower growth than the healthy reference)^5^. Subgrouping children based on up/down crossing of any LAZ threshold is particularly problematic when these subgroups are compared with respect to their linear growth trajectories. For example, LAZ-by-age trajectories shown in Figure 3b by HBGDki are compared across groups defined by the timing of stunting onset (a threshold-crossing definition)^1^. Yet, these trajectories cannot be interpreted because a child’s LAZ at one timepoint (used to define the subgroups) is mathematically coupled to the child’s prior and subsequent values^*^.

We further hypothesized that the dependence of stunting incidence/reversal estimates on the starting mean LAZ and between-timepoint correlations limits their usefulness as population-level indicators of LGF. To test this, we generated 25 simulated population datasets reflecting varying combinations of starting mean LAZ and LAZ changes between 0-3 months while holding between-timepoint correlation constant (**Extended Data Table 3**). These simulations demonstrate that the incidences of stunting onset and reversal are determined by the magnitude of the shift in mean LAZ and the mean LAZ at the start of the interval (**Figure 2a**). The effect of starting LAZ on both stunting onset and reversal raises concerns about using these indicators to compare the burden of LGF across populations with different baseline mean LAZ. For example, HBGDki’s observation of higher rates of earlier stunting onset (incidence at 0-3 months) in the SA cohorts compared to other regions of the world may be attributed to the lower mean LAZ at birth in SA^1^. Our simulations also show that stunting reversal is a statistical artifact that occurs in the absence of factors that promote the rapid growth of short children. Due to RTM, children selected from the lower tail of the distribution (i.e., LAZ <-2) are more likely, at the next measurement, to have LAZ values closer to the mean of the underlying population from which they were selected. Therefore, when the starting mean LAZ of a population is above -2, some stunted children will necessarily cross above the -2 threshold towards the population mean at subsequent measurements (i.e., reversal) simply due to the statistical phenomenon (Figure 2a). In fact, rates of reversal observed by HBGDki appear to be largely attributable to RTM (Figure 1).

**FIGURE 2.**
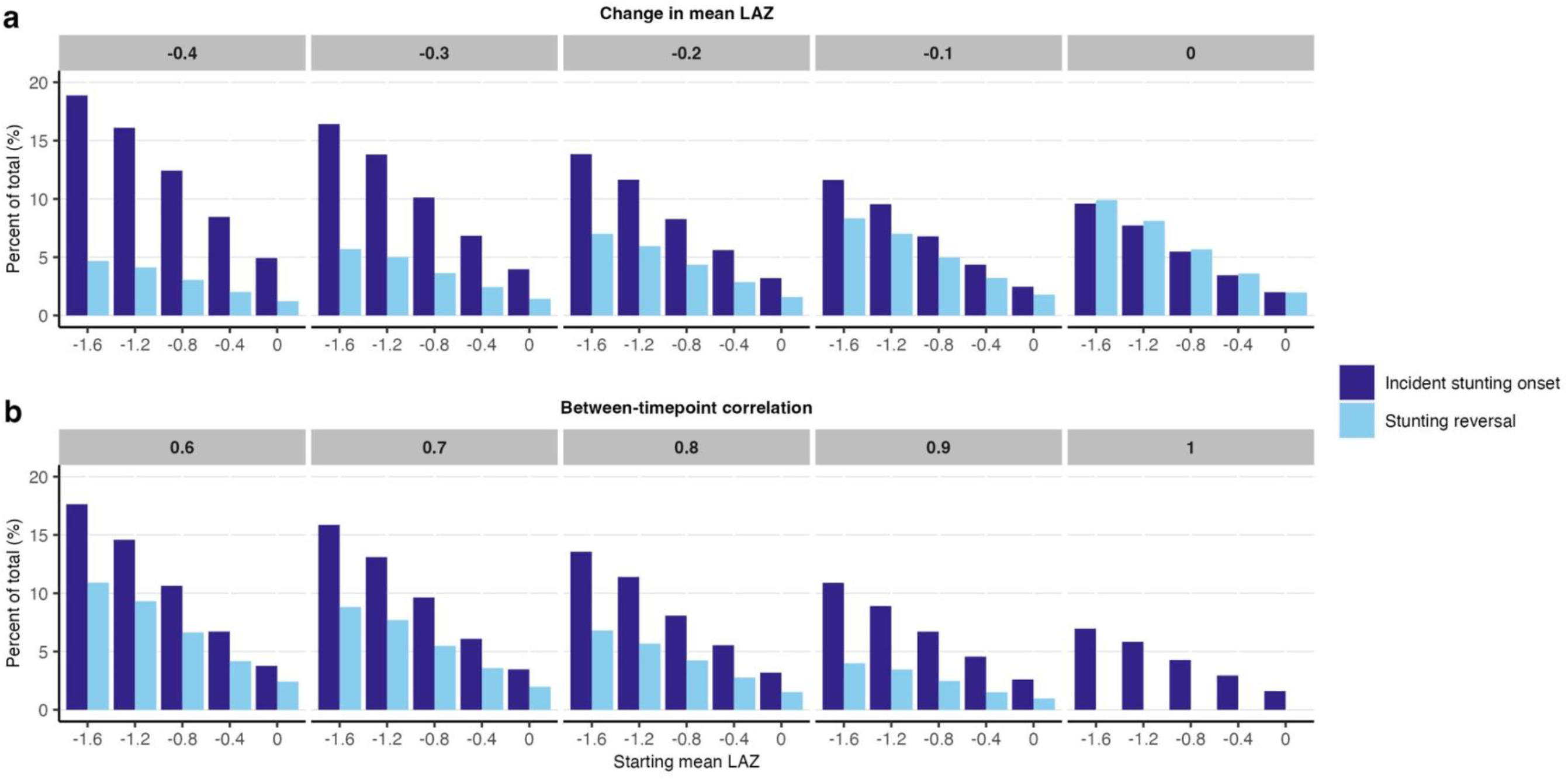
Incident stunting onset and reversal (%) by starting LAZ over a 3-month interval, for different magnitudes of a downward shift in the mean length-for-age z-score (LAZ) distribution (constant corr of 0.79) (Panel a) and increasing between-timepoint correlation (constant LAZ shift of -0.2) (Panel b). Lower starting mean LAZ or higher magnitude of downward LAZ distribution shifts increases the percentage of children with incident stunting onset (crossing below the stunting threshold of LAZ = -2) and decreases stunting reversal (crossing above LAZ = -2) (Panel a). Higher between-timepoint correlations decrease the percentage of children with incident stunting onset and stunting reversal, and this effect is more pronounced for stunting reversal (no reversal when corr = 1) (Panel b). Simulations and calculation of stunting incidence and reversal are explained in Supplementary Information.

**FIGURE 3.**
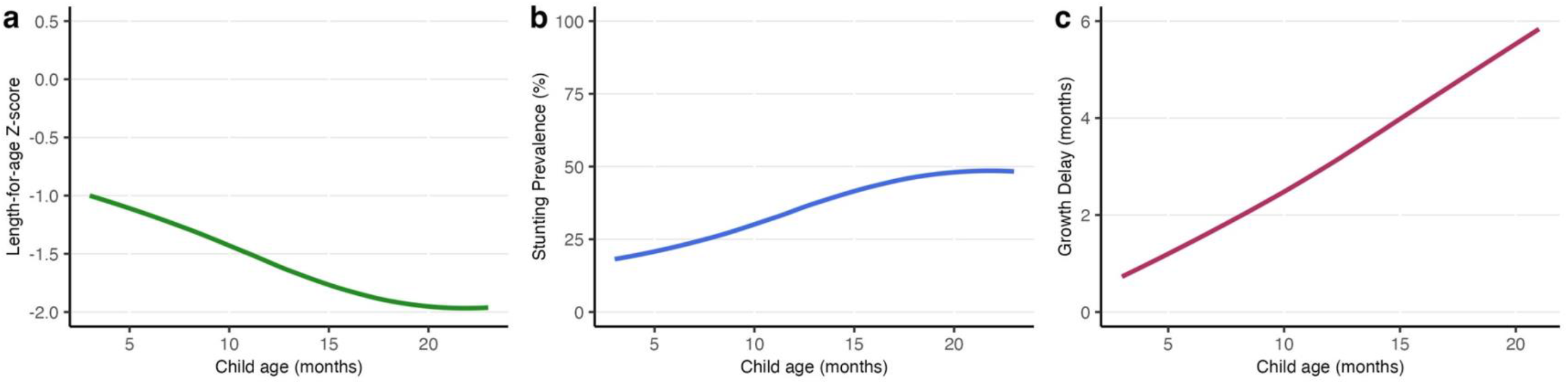
Mean length-for-age z-scores (LAZ) (Panel a), stunting prevalence (Panel b), and growth delay (Panel c) by age in a simulated cohort followed from 3 to 23 months of age. Declining LAZ-by-age trajectories as in panel (a) may be erroneously interpreted as showing that linear growth faltering primarily occurs in early infancy and that a disproportionately high proportion of the stunting burden is already present by 3 months of age (Panel b), corresponding to the inferences by Benjamin-Chung et al. However, when the same trajectory is expressed using growth delay (chronological age minus height-age) (Panel c), faltering is shown to proceed as a monotonic trend throughout and beyond infancy. A plateau at a negative mean LAZ, when mean LAZ and stunting prevalence are nearly constant with age (seen after ∼18 months in panels (a) and (b)), represents suboptimal growth for a population as demonstrated by the ongoing accrual of growth delay in panel (c). Simulations and calculation of growth delay are explained in Supplementary Information.

Next, we simulated 25 population datasets where the change in LAZ was held constant while the starting mean LAZ and between-timepoint correlation varied (Extended Data Table 3). These simulations show that stunting reversal rates were more strongly affected by changes in between-timepoint correlation than incident stunting onset across a range of starting mean LAZs (**Figure 2b & Extended Data Figure 1**). To further demonstrate how between-timepoint length/height correlation affects stunting incidence and reversal, we modified the simulated cohort used to generate the simulated incidence shown in Figure 1 by assuming a perfect correlation between timepoints (corr=1), effectively removing random error from the distribution. This demonstrated that the incidence of stunting in the simulated cohort was largely driven by the downward shift of the LAZ distribution, as threshold-crossing events occurred even in the perfect-correlation scenario (Figure 1). Conversely, stunting reversal requires an infant to move in the opposite direction of the group’s average trajectory (when the group is experiencing faltering) and, therefore, depends more on between-timepoint variation in LAZ (Figure 1). The strength of correlation between serial height/length measurements varies by age and duration of the time interval between measurements and can also be influenced by operational factors that are unrelated to children’s growth (e.g., using different length measuring instruments at different ages^7^). Therefore, the dependence of both indicators on between-timepoint correlation (particularly for reversal) undermines the validity of the between-age and between-region comparisons that supported the conclusions drawn from the HBGDki analysis.

Even if HBGDki had avoided subgrouping children based on threshold-crossing events, their conclusions would remain compromised by a reliance on age-related trajectories of mean LAZ and other LAZ-derived indicators. We previously demonstrated that LAZ tracking is poorly suited to quantifying the rate and timing of population-level LGF because it entails comparisons to healthy children of the same average *chronological* age and unjustifiably considers population tracking along negative z-scores to be normal^8^. Faltering due to suboptimal environmental and nutritional conditions uncouples chronological age from skeletal maturity, such that the potential linear growth of a population during an observed interval cannot be predicted by chronological age^9^. Alternative approaches using height-age (i.e., the age at which the observed mean height of a population would be considered normal/healthy) and growth delay (i.e., the difference between height-age and the chronological age) provide representations of faltering and catch-up that are more consistent with biological mechanisms of long bone growth and the theory of growth plate senescence^10^. We previously showed that in contrast to LAZ-by-age slopes, growth delay-age slopes correlate more strongly and coherently with population indicators such as child mortality^11^. Use of height-age and growth delay counter the assertion that LGF disproportionately occurs in early infancy, instead demonstrating that LGF in LMICs proceeds unrelentingly throughout infancy and beyond (**Figure 3**), and that LGF may be ongoing even when mean LAZ trajectories misleadingly suggest a plateau (**Figure 3**) or recovery (**Figure 4**).

**FIGURE 4.**
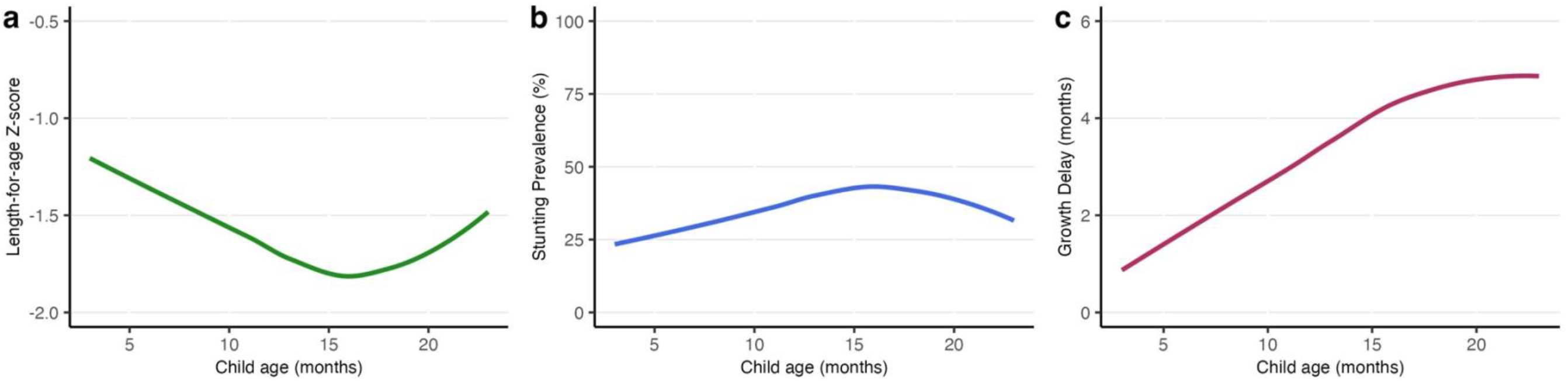
Mean length-for-age z-score (LAZ) (Panel a), stunting prevalence (Panel b), and growth delay (Panel c) by age in a simulated cohort that experiences an initial decline in mean LAZ followed by an increase in mean LAZ from 3 to 23 months of age. Increases in mean LAZ towards zero (Panel a) and declining stunting prevalence (Panel b) after ∼16 months of age may be mistakenly interpreted as ‘catch-up’ growth, even though they can be observed in the context of ongoing accrual of growth delay (Panel c). Simulations and calculation of growth delay are explained in Supplementary Information.

We have outlined three limitations of the metrics and methods used in the HBGDki investigators’ analyses: (i) incident stunting onset and reversal are artifacts of whole-population shifts in the height distribution in LMICs rather than meaningful representations of individual children’s growth patterns; (ii) stunting incidence/reversal frequencies are ill-suited for between-age or between-population comparisons because they are affected by between-timepoint correlations (and hence, RTM), the magnitudes of which differ by age and setting; and, (iii) rather than LAZ-tracking with age used in the HBGDki analyses, height-age and growth delay more appropriately represent population-average growth patterns, revealing that faltering starts early and proceeds throughout the first two years and beyond, rather than occurring primarily in the first few months. HBGDki’s adoption of LAZ-tracking and threshold-crossing indicators (stunting incidence or reversal) reflects the conventional and widespread use of stunting in epidemiological research^3^. However, we believe this approach leads to incorrect conclusions about the timing of faltering and distracts from efforts to understand why nearly *all* children in most LMICs grow more slowly than expected for their chronological age. The undue emphasis on stunting in this field is comparable to seeing a sinking ocean liner and believing that only people on the lower decks need lifeboats. In future studies of child growth in LMICs, the adoption of approaches that reflect whole-population shifts in height distributions and use of height-age (rather than LAZ/stunting) will more effectively support public health actions that benefit all children throughout infancy and beyond.

## Data Availability

The code used to generate the simulated datasets used in this analysis is available at https://github.com/DiegoGBassani/Linear_growth_faltering.

## Supplementary Information

### Data sources

We used simulated datasets to calculate incident stunting onset and stunting reversal shown in Figures 1 and 2, and stunting prevalence shown in Figures 3 and 4. While the simulated datasets used in Figures 1 to 4 differ based on the specified mean LAZ values for a given time point and the age intervals, all simulations involved the following general steps:

1. Set the parameters.
  a. Define the age range.
  b. Define mean LAZ for each follow-up timepoint (e.g., mean LAZ at 3-months, 6-months, etc.).
  c. Define the standard deviation (SD) for each specified mean LAZ. We held the SD constant at 1.1 (a value chosen based on Benjamin-Chung et al.’s Extended Data Figure 7) for all datasets to simulate whole population shifting.
  d. Define the correlation structure between LAZ measurements. We based between-timepoint correlations on a Bangladeshi birth cohort with high-quality anthropometric data collected tri-monthly from birth to 24 months^12^; the predicted correlations (used in our simulations)^†,‡^ between the intervals presented in Figures 1-4 are available in Extended Data Tables 1 and 3, and the full correlation matrices are available online^13^.
  e. Set the number of observations of the simulated dataset. We chose N=10,000 for all datasets.
2. Simulate the cohort. We assumed a normal LAZ distribution for each dataset, and simulated 10,000 observations adhering to the mean LAZ, SD, and correlation structure defined in Step 1. We used the *drawnorm* command in Stata – for example, “drawnorm laz0 laz3 laz6, means(Insert Defined Means) sds(Insert Defined SDs) corr(Insert Defined Correlation Structure)”.
3. Calculate incident stunting onset, stunting reversal, and/or stunting prevalence in the simulated dataset.

### Figure 1

#### Incident stunting onset estimates

A cohort dataset was simulated as described above (normal LAZ distribution; constant LAZ SD of 1.1; N=10,000) to calculate incident stunting onset estimates to compare with those reported in Benjamin-Chung et al.’s Figure 3(a) for the South Asian (SA) cohort (shown as “reported incidence” in Figure 1). The mean LAZ values corresponding to the SA cohort’s estimates in Figure 3(a) were extracted tri-monthly (from 0-24 months, e.g., mean LAZ at 3 months, at 6 months, at 9 months (…) 24 months) from Benjamin-Chung et al.’s Supplementary Information Figure 3.1.1, depicting mean LAZ-by-age for their SA cohorts, using the online tool Plot Digitizer (plotdigitizer.com/app). As Figure 3.1.1 only showed the mean LAZ starting at 3 months, we assumed the mean LAZ at 0 months was -1.0. The extracted mean LAZ values and correlation coefficients used in this simulation are reported in Extended Data Table 1. Stunting incident onset estimates calculated from this simulated dataset are referred to as “simulated incidence” in Figure 1. This simulation was then repeated but using a tri-monthly correlation matrix that assumed perfect between-timepoint correlations (corr=1), denoted “simulated incidence (corr=1)” in Figure 1.

#### Stunting reversal estimates

To calculate the stunting reversal estimates shown in Figure 1, we simulated a cohort dataset with monthly LAZ measurements (from 0 to 15 months) using parameters previously described (normal LAZ distribution; constant LAZ SD of 1.1; N=10,000). Monthly measurements were selected for comparability with Benjamin-Chung et al.’s Extended Data Figure 12. We used Plot Digitizer to extract reversal estimates for the SA cohort from this figure, and they are presented in Figure 1 as “reported reversal”. The mean LAZ values corresponding to Benjamin-Chung et al.’s reported reversal estimates were extracted from their Supplementary Information Figure 3.1.2 at monthly intervals using Plot Digitizer. The extracted mean LAZ values and correlation coefficients used in this simulation are presented in Extended Data Table 1. Stunting reversal estimates calculated from this simulated dataset are referred to as “simulated reversal” in Figure 1. This simulation was also repeated, using a monthly correlation matrix that assumed perfect between-timepoint correlations (corr=1), denoted “simulated reversal (corr=1)” in Figure 1.

The purpose of calculating incident stunting onset estimates using a simulated dataset with perfect between-timepoint correlations among repeated LAZ values (corr=1 for all intervals) was to show, in each interval, the proportion of infants who experienced their first episode of stunting (among all infants) that would result entirely from the shift in the LAZ distribution in that interval. Perfect between-timepoint correlation means that all individual children experience the same magnitude of change in LAZ in each interval and retain their rank within the population. The difference between each “simulated incidence (corr=1)” and the corresponding “simulated incidence” estimate in Figure 1 can be interpreted as the percentage-point increment in the cumulative incidence of new-onset stunting due to within-child variations in growth patterns. As with incident stunting onset, the purpose of generating stunting reversal estimates in a simulated population dataset with perfect between-timepoint correlations is to show, in each interval, the proportion of infants who experienced reversal (among all infants) entirely due to the shift in the LAZ distribution in that interval, since the perfect between-timepoint correlation eliminates regression to the mean (RTM). The absence of reversal in this scenario highlights that when the LAZ distribution is shifting down, all episodes of reversal can be attributed to RTM.

Simulations were implemented using Stata 17. Correlation coefficients used in the simulated datasets for Figure 1 are available in Extended Data Table 1 (showing only the between-timepoint correlations reflecting the intervals presented in Figure 1), and the complete correlation matrices are available in our online, publicly available code^13^. The Figure 1 scatterplot was generated using R, where circles were used to denote point estimates and loess curves (span/bandwidth = 0.75) were used to smooth across each set of points.

### Figure 2

For Figure 2(a), we simulated 25 population datasets from which incident stunting onset and reversal estimates were generated over the 0–3-month interval only. Datasets were simulated using parameters described earlier (normal LAZ distribution; LAZ SD of 1.1; N=10,000), with the combination of the 0- and 3-month mean LAZs differing for each dataset. Specifically, we introduced variation in the starting position and magnitude of the whole-population distribution shift by altering the starting mean LAZ (5 discrete values: 0, -0.4, -0.8, -1.2, and -1.6) and the change in mean LAZ from 0- to 3-months (5 shifts per starting LAZ: 0, -0.1, -0.2, -0.3, and -0.4), such that each cohort dataset reflected a unique combination of mean LAZ values at baseline (birth) and endline (3 months) (Extended Data Table 3). The between-timepoint correlation was held constant (corr=0.79), corresponding to the predicted between-timepoint correlation for the 0-to 3-month measurements from the control group of the Bangladeshi trial cohort.

For Figure 2(b), we simulated 25 population datasets to calculate onset and reversal estimates over the 0–3-month period, as was done for Figure 2(a), but instead, the change in LAZ was held constant at -0.2, and the between-timepoint correlation was varied (corr=0.6, 0.7, 0.8, 0.9, and 1.0) (Extended Data Table 3). The same mean starting LAZ values were used as in Figure 2(a), such that there were five unique combinations of starting (0-month) and 3-month LAZ in the cohort datasets, as the changing correlation presented the main source of variation across all datasets.

### Figures 3 and 4

To calculate stunting prevalence estimates for Figure 3, we generated a simulated dataset using the mean LAZ values extracted at monthly intervals from the mean LAZ-by-age trajectory for the SA cohort reported in Benjamin-Chung et al.’s Supplementary Information Figure 3.1.2 for ages 3-23 months (Extended Data Table 1). Conversely, for the dataset simulated for Figure 4, we deliberately chose mean LAZ values at monthly intervals for ages 3-23 months to illustrate an increase in the LAZ-by-age trajectory (Extended Data Table 1). Otherwise, for both Figures 3 and 4, simulations were performed as described above – assuming whole-population shifting (constant SD=1.1), normal LAZ distribution, 10,000 observations at each age, and realistic between-timepoint correlations (reported in Extended Data Table 1 and full matrices available from our code online^13^). Growth delay was calculated from the simulated mean LAZ (nearly identical to the input mean LAZ values shown in Extended Data Table 1), as further described in the “Calculating Growth Delay” section.

For Figures 2, 3, and 4, simulations were conducted using Stata 17 and plots for all figures were generated using R, and for which the code is available online^13^.

### Calculating and interpreting linear growth faltering indicators

**‘Incident stunting onset’**, also referred to by Benjamin-Chung et al. as ‘newly stunted’, was calculated using the method of Benjamin-Chung et al., whereby each estimate reflects the proportion of all children (N=10,000) measured at one timepoint who had LAZ<-2 (stunting) for the first time (children whose LAZ was previously always >=-2 and with LAZ <-2 at the current age). As ‘incident stunting onset’ is based on each infant’s first episode, it is closely related to the cumulative incidence function and specifically reflects the percentage-point increment in cumulative incidence in each successive age interval; however, ‘incident stunting onset’ frequencies expressed as proportions of all infants do not represent true incidence proportions for each interval since the population included in the denominator was not restricted to infants who were at-risk of stunting at the start of each interval.

**‘Stunting reversal’** (also referred to by Benjamin-Chung et al. as ‘stunting reversed’) was calculated using the method of Benjamin-Chung et al., whereby each estimate reflects the proportion of children in the current interval with LAZ>=-2 (‘not stunted’) who had LAZ<-2 in the preceding interval, among all children measured in the current interval (N=10,000). As with incident stunting onset, reversal frequencies expressed as proportions of all infants do not represent true incidence proportions for each interval since the population included in the denominator was not restricted to infants who were stunted at the start of each interval.

In these simulated cohorts, we assumed no missingness, loss-to-follow-up, or death, such that the sample size (N=10,000) was the same at each measurement. While sample sizes in Benjamin-Chung et al.’s real-world dataset varied by interval, it remained unclear how they classified infants measured in one interval but with missing data from prior intervals; for example, if infants identified as stunted in the 0-3 month interval were not measured at birth, the assumption that these infants were ‘newly stunted’ at 0-3 months would bias estimates of incident stunting onset upwards in that interval (Extended Data Table 2). Therefore, missingness patterns and imputation decisions likely account for some of the differences between the observed and simulated populations.

### Calculating growth delay

Growth delay was calculated as described in Mansukoski et al.^8^ Briefly, mean LAZ was generated at monthly intervals (3-23 months) from the simulated population datasets used for Figures 3 and 4, and mean chronological age was assumed to be equal to the time of measurement, in days, using a conversion factor of 1-month equals 30.4375 days. Mean height-age was determined from mean LAZ, by finding the age corresponding to the WHO Growth Standards (WHO-GS) median length which most closely corresponded to the mean length back-calculated from mean LAZ. The WHO-GS median lengths are available in the ‘lenanthro.dta’ file from https://github.com/unicef-drp/igrowup_update; this data file was collapsed by sex, i.e., taking a simple average of the parameters presented separately for females and males. Mean growth delay was calculated as mean chronological age minus mean height-age. The STATA code is available online^13^.

## Acknowledgements

We thank N. Perumal for providing constructive feedback in the development of this paper, C. Y. Chen for independent verification of the simulations, and H. Qamar for verification of the height-age and growth delay calculations.

## Extended data tables and figures

**Extended Data Figure 1.**
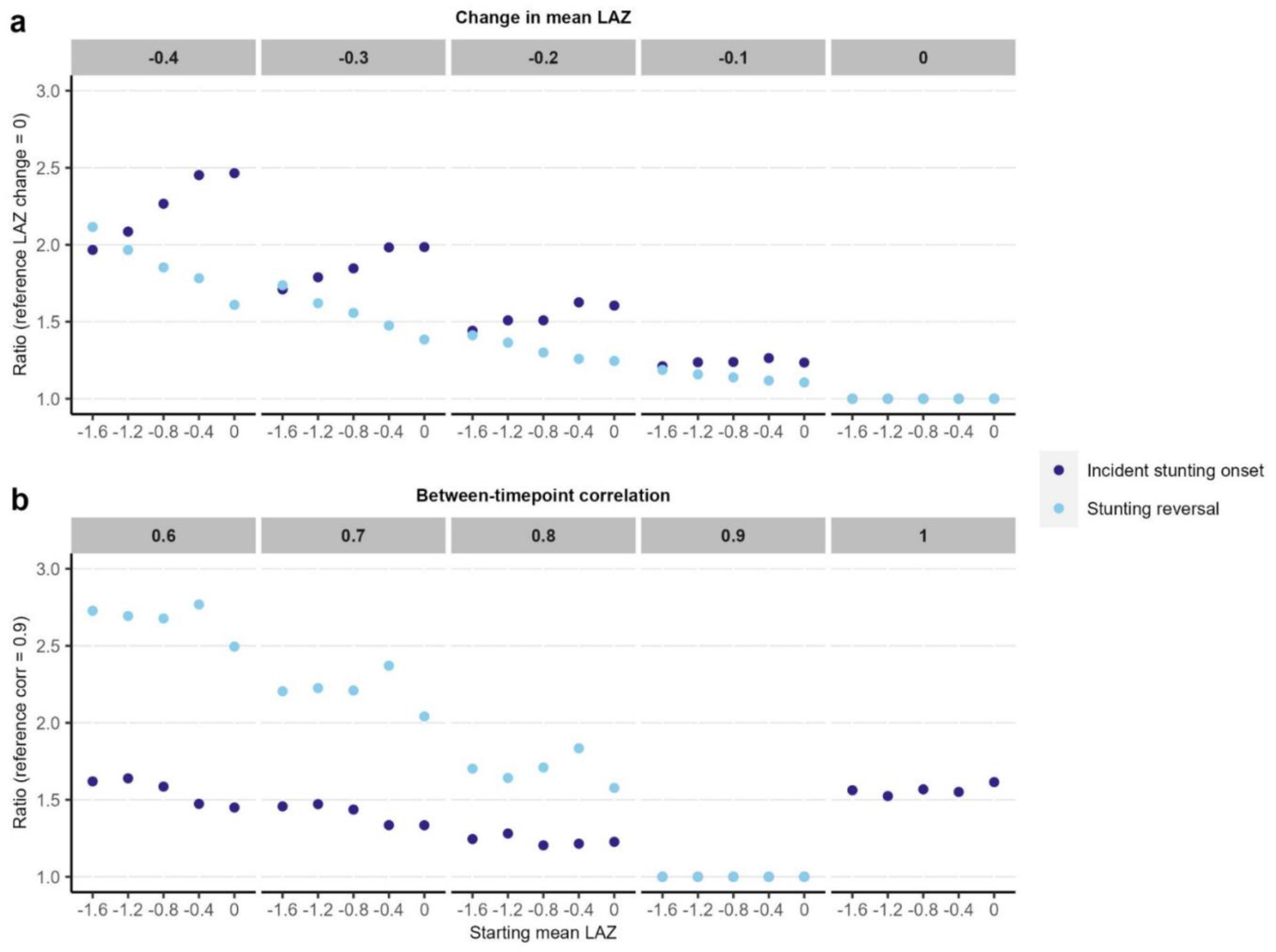
Re-expression of data shown in main Figure 2 using ratios to compare incident stunting onset and reversal rates in each scenario to a corresponding reference scenario. The reference change in mean LAZ was 0 (Panel a), and the reference correlation coefficient was 0.9 (Panel b). Where the observed value was greater than the reference value, ratios were calculated with the reference value in the denominator, and where the observed value was less than the reference value, the ratio was inverted, such that a two-fold increase is presented instead of a reduction by half. This approach was used to facilitate comparisons between incident onset and reversal estimates in panel.

**Extended Data Table 1.**
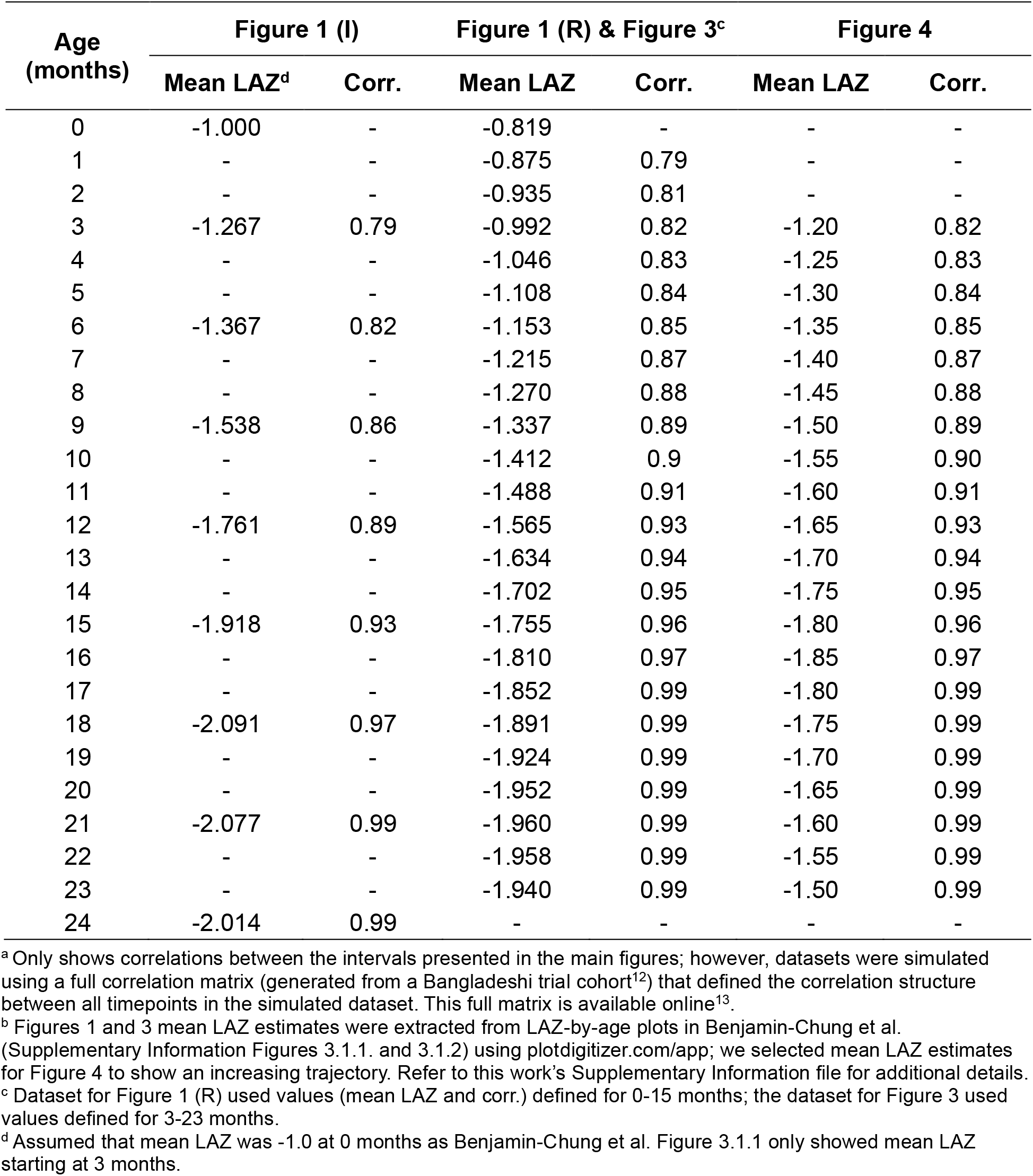
Mean length-for-age z-scores (LAZ) and correlation coefficients (corr.) used to generate simulated datasets from which incident stunting onset (I), stunting reversal (R), and stunting prevalence were calculated in Figures 1, 3, and 4 ^a, b^

**Extended Data Table 2.** Incident stunting onset at 0 and 3 months of age in a simulated cohort with realistic between-timepoint correlation and assuming 10% and 20% missingness at birth ^a^.

| Age interval, months | Mean LAZ | Missingness at birth | Sample size | Incident stunting onset, count (%) |  |
| --- | --- | --- | --- | --- | --- |
|  |  |  |  | With missingness<sup>b</sup> | No missingness<sup>c</sup> |
| 0 | -1.0 | 10% | 9000 | 1648 (18.3) | 1833 (18.3) |
| >0-3 | -1.3 | 10% | 10000 | 1283 (12.8) | 1153 (11.5) |
| 0 | -1.0 | 20% | 8000 | 1467 (18.3) | 1833 (18.3) |
| >0-3 | -1.3 | 20% | 10000 | 1418 (14.2) | 1153 (11.5) |

LAZ, length-for-age z-score.
<sup>a</sup> The simulated cohort dataset for “Simulated incidence” estimates in Figure 1 was used, assuming 10% and 20% missingness at birth; correlation in LAZ between 0-3 months was 0.79, selected from a correlation matrix generated from a Bangladeshi trial cohort<sup>12</sup>; the code for this simulation is available online<sup>13</sup>.
<sup>b</sup> Infants with missing birth measurements were considered ‘newly stunted’ if their 3-month LAZ <-2.
<sup>c</sup> Values are Figure 1 ‘Simulated incidence’ estimates, displayed for comparison.

**Extended Data Table 3.**
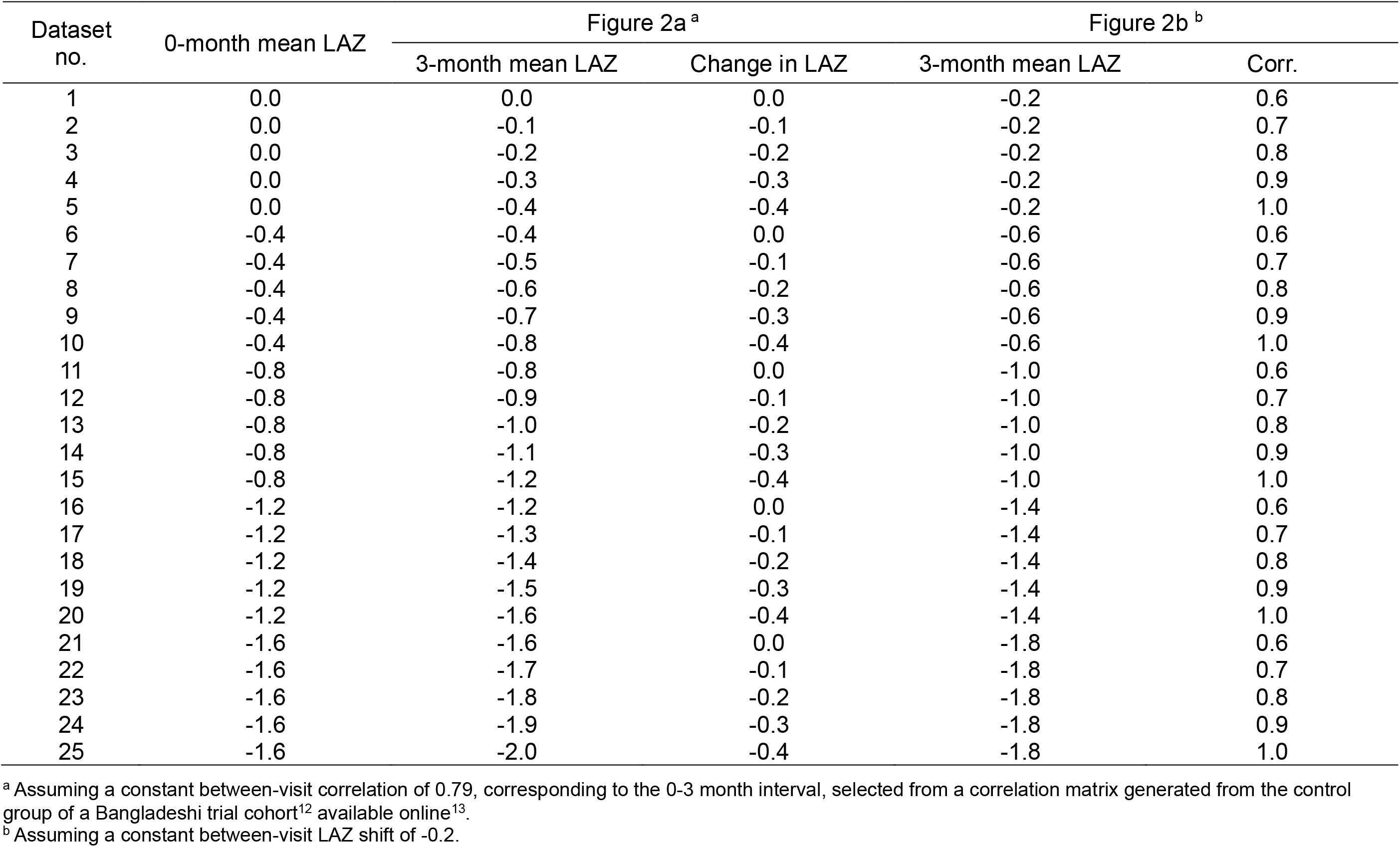
Starting mean length-for-age z-score (LAZ) (0-months), 3-month mean LAZ, change in mean LAZ (‘LAZ distribution shift’) and correlation coefficients (corr.) used to simulate population datasets used in Figure 2a and 2b.

## Footnotes

* A similar point was made during peer review of the HBGDki manuscript, where a reviewer referred to this relationship as tautological^6^.

† In our prior unpublished work, we developed a model to estimate monthly correlations using the correlation values generated from the tri-monthly length measurements in this Bangladeshi trial cohort (placebo group only); predicted correlations from the resultant model were used in this study.

‡ We acknowledge that the correlations used in our simulations do not reflect the actual between-timepoint correlations for Benjamin-Chung et al.’s South Asian cohort, contributing variation between their real-world and our simulated populations.

